# HeartNet: Self Multi-Head Attention Mechanism via Convolutional Network with Adversarial Data Synthesis for ECG-based Arrhythmia Classification

**DOI:** 10.1101/2021.12.20.21268090

**Authors:** Taki Hasan Rafi, Young Woong-Ko

## Abstract

Cardiovascular disease is now one of the leading causes of morbidity and mortality in humans. Electrocardiogram (ECG) is a reliable tool for monitoring the health of the cardiovascular system. Currently, there has been a lot of focus on accurately categorizing heartbeats. There is a high demand on automatic ECG classification systems to assist medical professionals. In this paper we proposed a new deep learning method called *HeartNet* for developing an automatic ECG classifier. The proposed deep learning method is compressed by multi-head attention mechanism on top of CNN model. The main challenge of insufficient data label is solved by adversarial data synthesis adopting generative adversarial network (GAN) with generating additional training samples. It drastically improves the overall performance of the proposed method by 5-10% on each insufficient data label category. We evaluated our proposed method utilizing MIT-BIH dataset. Our proposed method has shown 99.67 ± 0.11 accuracy and 89.24 ± 1.71 MCC trained with adversarial data synthesized dataset. However, we have also utilized two individual datasets such as Atrial Fibrillation Detection Database and PTB Diagnostic Database to see the performance of our proposed model on ECG classification. The effectiveness and robustness of proposed method are validated by extensive experiments, comparison and analysis. Later on, we also highlighted some limitations of this work.

## 1 Introduction

Cardiovascular disease (CVD) is the leading cause of mortality in humans, accounting for 31% of all deaths globally in 2019, 85 percent occurred as a result of a heart attack. Diagnosis of cardiovascular disease is based on patient’s clinical examinations and current stage of their cardiovascular system. Due to the huge quantity of heterogeneous data that must be dealt with in many situations, the conventional rule-based diagnostic paradigm is inefficient and needs substantial analysis and medical knowledge to attain sufficient accuracy in diagnosis [1]. Furthermore, since health monitoring systems are extremely sensitive to anomalies in the ECG, which can prevent them from missing severe cardiovascular events, a large fraction of ECG recordings are typically false alarms. As a result, there is an increasing demand for physicians’ assistance in interpreting ECG records [2].

The ECG images are manually interpreted by medical professionals to diagnose the patient’s cardiac condition. With the advancement of state-of-art technologies, numerous automated diagnostic techniques for the diagnosis and detection of arrhythmias have been created to aid clinicians. ECGs are extensively used to diagnose and classify arrhythmias [3]. Whenever a whole ECG signal of a patient is not accessible and only numerous single heartbeats or a limited part of the ECG signals are available, the findings of heartbeat categorization will be utilized to assess the present patient’s heart disease [4]. A Holter monitor and a conventional 12-lead configuration comprising of three limb leads, three pressured limb leads, and six thoracic leads are used to get an Electrocardiogram report [5]. With the advancement of Artificial Intelligence, numerous machine learning approaches are being utilized in the classification of ECG signal features, with the goal of resolving issues such as vast quantities of ECG signal feature data [6]. Because of numerous contemporary medical applications where such problem may be mentioned, the relevance of ECG classification is currently quite high. However, the usage of heuristic hand-crafted or manufactured features with shallower feature learning frameworks is one of the primary drawbacks of these ML solutions [7, 8].

Deep learning has recently been identified as a viable option for ECG interpretation. Deep convolutional neural networks extract features from raw input images or signals automatically. Deep neural networks-based ECG interpretation is becoming increasingly attractive due to its excellent performance in automated categorization [12].

Deep learning has shown enormous advances on health-care. Deep neural network algorithms like CNN, LSTMs and other hybrid models are extensively used in various tasks like image classification, text mining, signal classification and disease detection [13]. To increase the performance of classification models, it is needed to have an automatic robust feature extraction. Which can take key features to classify ECG signals. There are some challenges that are required address to while developing a deep learning based arrhythmia classifier. Key challenges are **(1)** feature selection manually, **(2)** developed techniques that are utilized for feature extraction, and **(3)** developed deep learning models that are utilized for classification on imbalanced dataset. Domain knowledge is required for automatic feature extraction from ECG images.

Our main aim is to address and solve the insufficient data label problem. Handling classes with insufficient data label is difficult for a deep learning classifier. The primary goal of this work is to thrive a new arrhythmia classification model based on deep learning. So We have developed an deep learning arrhythmia classifier utilizing multi-head self attention mechanism with convolutional network. We worked with MIT-BTH dataset in our experiment, but initially the dataset is imbalanced. So we adopted generative adversarial network to increase the number of data in the classes with insufficient data labels. However, the proposed model has performed well in each category classification task. Futhermore, we tested two other datasets to ensure validation of our proposed model. The summary of our contributions are following:

- We proposed a new high-performing ECG classifier called *HeartNet*, combining multi-head attention mechanism on top of convolutional network.
- We utilized generative adversarial network (GAN) to synthesized insufficient labeled category. It increases the number of sample data on each insufficient data label category and improves overall performance on each category of our proposed model.
- Two individual heart disease detection datasets are being utilized to see the effectiveness and generalization of our proposed model in various conditions.
- We reviewed and demonstrated a recent overview of deep learning techniques on arrhythmia classification and compared recent outcomes with our proposed model.

The reminder part of our paper is organized into five sections. Section II presents the related works on arrhythmia classification. Section III introduces the overview of methods used in this experiment. In Section IV, description of the experimental dataset, all experimental results and simulations are provided with a comprehensive comparison. And finally, in the Section V presents the limitations of this work and Section VI presents the conclusion of this whole outcome.

## 2 Related Works

The section focuses on summarizing and comparing the notable studies within 2017 to 2021 based on the deep learning based models to cope up with the arrhythmia classification problem. We come across several recent state-of-art deep learning based arrhythmia classification works and present a holistic overview of recent works on arrhythmia classification models with comparing their performances. We also tried to depict the recent effective arrhythmia classification techniques together with indicated accuracy by analyzing the literature. This would enable researchers for additional analysis to their demands.Previously arrhythmia classification on ECG signals classifications were mainly focused on signal processing techniques. However recent approaches have focused on deep learning.

Weimann and Conrad [2] has utilized transfer learning in their experiment. They mainly trained CNN model on raw ECG data, then took a small sub-set of the dataset to classify atrial fibrillation. They fine-tuned the CNN model to reduce the number parameters and improve the performance of the proposed model. Wu et al. [4] proposed an efficient 1D-CNN framework consisting of 12 layers for arrhythmia classification. They also adopted a denoising method with the wavelet self-adoption feature. Niu et al. [5] proposed a new deep-learning framework with adversarial domain adaptation for ECG classification. With the help of domain adaptation they increased the lower data samples to improve the effectiveness of their model. Liang et al. [6] proposed a new deep learning framework with a combination of CNN and bidirectional LSTM models. They also utilized evolutionary neural network to compare with their proposed model. Abdullah and Ani [7] proposed a 1D CNN and long short term memory (LSTM) based framework for ECG classification. They utilized two extensively common databases in their validation. Abdalla et al. [8] adopted a convoluational neural network consisting of 11 layers for 10-class arrhythmia classification. They distributed these 11 layers into several parts to achieve higher accuracy. Kim and Pyun [10] proposed a bidirectional long-short term memory network based on RNN for ECG classification. They also derived data preprocessing with normalization and derivative filtering. Alarsan and Alarsan [12] adopted various machine learning algorithms on arrhythmia classification. They implemented these machine learning models with MLlib library. Where random forest classifier has achieved higher accuracy in multi-class problem. Zhang et al. [13] proposed a deep-CNN model. They used time domain transformation to convert 1D signals to 2D images to feed their proposed model. They also adaopted band pass filtering method to extract features from ECG raw data. Liu et al. [14] proposed an attention based hybrid CNN-LSTM model for arrhythmia classification. The LSTM model is designed with stacked LSTM architecture and the CNN model is designed with 2D architecture. Saadatnejad et al. [15] proposed a novel deep learning model with multiple LSTM-RNN networks. The proposed model also has wavelet transformation feature which made this model more effective and lightweight. Kachuee et al. [16] utilized a deep-convolutional neural network model with residual block to predict arrhythmia and myocardial infection. They used two databases in their experiment.

Zhang et al. [17] adopted a 1D convolutional neural network model with global pooling average to classify arrhythmia efficiently. Data pre-processing is also utilized in their experiment. Li et al. [18] proposed a 2D convolutional network with a feature of adaptive learning rate with Adadelta and biased dropout strategy. The proposed model anticipated with a higher accuracy and precision. Mattila et al. [19] adopted a convolutional neural network for inter-patient arrhythmia classification. They exploited various filtering and normalization techniques in the data preprocessing stage. Rahhal et al. [20] adopted a pre-trained VGGNet model in their ECG classification task. They used continuous wavelet transform and converted the signals into time domain. They additionally applied pre-trained to extract features before training their proposed model.

Mathews et al. [21] utilized restricted boltzmann machine and deep belief network in their experiment. Their proposed model has performed well in the lower sampling condition with simple features. Singh et al. [22] adopted a recurrent neural network, LSTM for ECG classification. They compared their proposed model with RNN and gated recurrent unit. Yildirim [23] proposed a novel deep learning ECG classifier with bidirectional LSTM. They introduced a wevelet sequence based layer in their model to generate new ECG signal sequences. They also developed undirectional (ULSTM) model to compare with the bidirectional (BiLSTM) model. Pyakillya et al. [24] adopted a 1D-CNN model with consists of 7 layers. Also the adopted model consists of global pooling average, three fully-connected layers, dropout and softmax layer which has 4 outputs. Li et al. [25] also adapted a 1D-CNN architecture which consists of 5 layers with 2 layers in downsampling and two layers for convolution.

In fact, there are many deep learning based methods are utilized in recent years for ECG classification. But there are still few methods are still not compatible with ECG signal classification. There are some limitations and shortcomings with these approaches. So tried to address them and build a robust solution which can assist medical professionals in heart problem diagnosis. In Table 1, a summary of our literature review is provided.

**Table 1:**
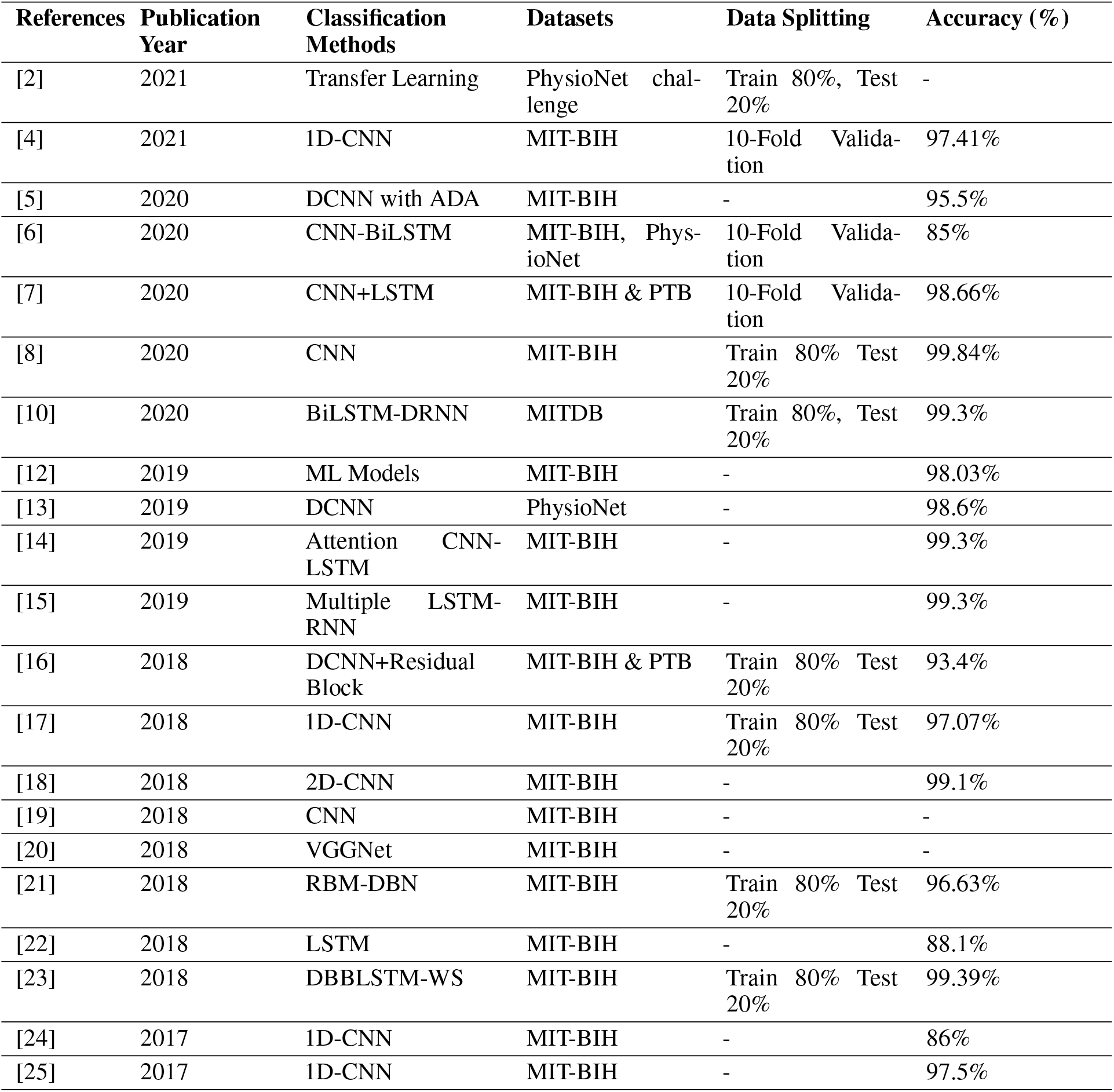
Recent Arrhythmia Classification Techniques Summary.

| References | Publication Year | Classification Methods | Datasets | Data Splitting | Accuracy (%) |
| --- | --- | --- | --- | --- | --- |
| [2] | 2021 | Transfer Learning | PhysioNet challenge | Train 80%, Test 20% | - |
| [4] | 2021 | 1D-CNN | MIT-BIH | 10-Fold Validation | 97.41% |
| [5] | 2020 | DCNN with ADA | MIT-BIH | - | 95.5% |
| [6] | 2020 | CNN-BiLSTM | MIT-BIH, PhysioNet | 10-Fold Validation | 85% |
| [7] | 2020 | CNN+LSTM | MIT-BIH & PTB | 10-Fold Validation | 98.66% |
| [8] | 2020 | CNN | MIT-BIH | Train 80% Test 20% | 99.84% |
| [10] | 2020 | BiLSTM-DRNN | MITDB | Train 80%, Test 20% | 99.3% |
| [12] | 2019 | ML Models | MIT-BIH | - | 98.03% |
| [13] | 2019 | DCNN | PhysioNet | - | 98.6% |
| [14] | 2019 | Attention CNN-LSTM | MIT-BIH | - | 99.3% |
| [15] | 2019 | Multiple LSTM-RNN | MIT-BIH | - | 99.3% |
| [16] | 2018 | DCNN+Residual Block | MIT-BIH & PTB | Train 80% Test 20% | 93.4% |
| [17] | 2018 | 1D-CNN | MIT-BIH | Train 80% Test 20% | 97.07% |
| [18] | 2018 | 2D-CNN | MIT-BIH | - | 99.1% |
| [19] | 2018 | CNN | MIT-BIH | - | - |
| [20] | 2018 | VGGNet | MIT-BIH | - | - |
| [21] | 2018 | RBM-DBN | MIT-BIH | Train 80% Test 20% | 96.63% |
| [22] | 2018 | LSTM | MIT-BIH | - | 88.1% |
| [23] | 2018 | DBBLSTM-WS | MIT-BIH | Train 80% Test 20% | 99.39% |
| [24] | 2017 | 1D-CNN | MIT-BIH | - | 86% |
| [25] | 2017 | 1D-CNN | MIT-BIH | - | 97.5% |

## 3 Methods

### 3.1 Generative Adversarial Network

Generative adversarial networks (GANs) are gaining popularity among deep learning researchers. GANs have been applied to various domains such as computer vision, natural language processing, data synthesis, semantic segmentation and other related areas. Multi-layer perceptrons are the straightforward implementing rules for adversarial modeling. The architecture of a typical GAN is shown in Figure 1. There are two essential parts in the GAN the architecture. The first one is Discriminator (*D*) which mainly disseminate between real images and generated images. However, another part is Generator (*G*) which generates fake images. The discriminator network is a convolutional network that can classify the examples supplied to it, a binomial classifier that labels cases as real or fake. In a sense, the generator is an inverted convolutional network: whereas a conventional convolutional classifier down-samples an example to produce a probability, the generator up-samples a vector of random noise to produce a probability. As an example The first dissipates data by using down-sampling techniques. The first generates new data, similar to maximum pooling, and the second generates new data.

**Figure 1:**
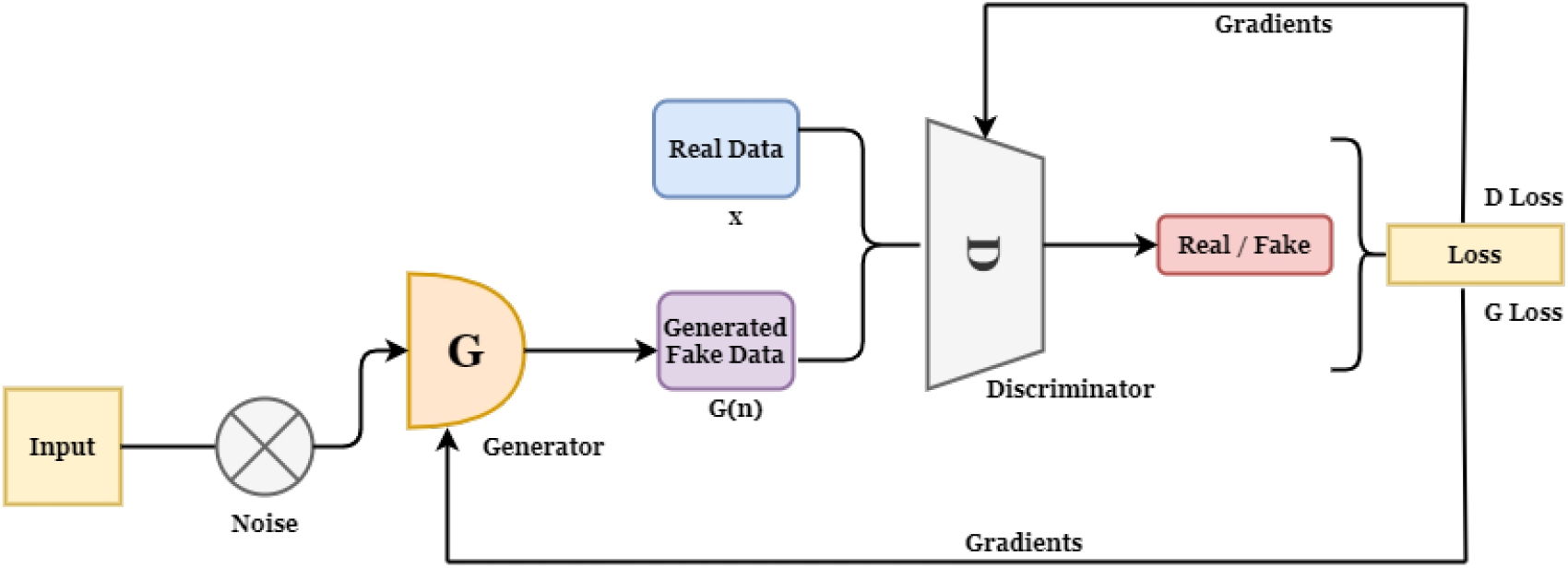
Generative Adversarial Network Architecture.

**Figure 2:**
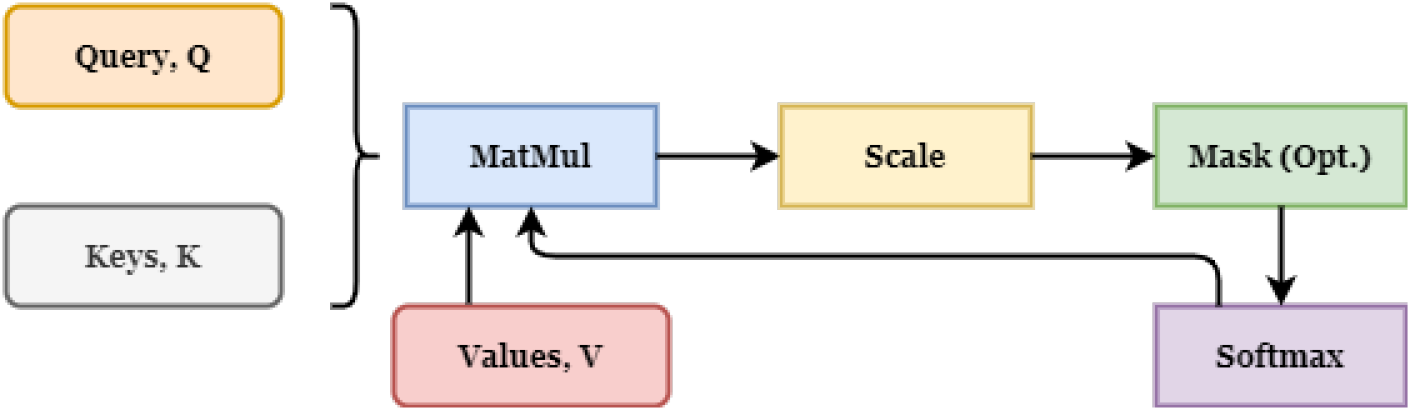
Scaled Dot Product Based Attention.

**Figure 3:**
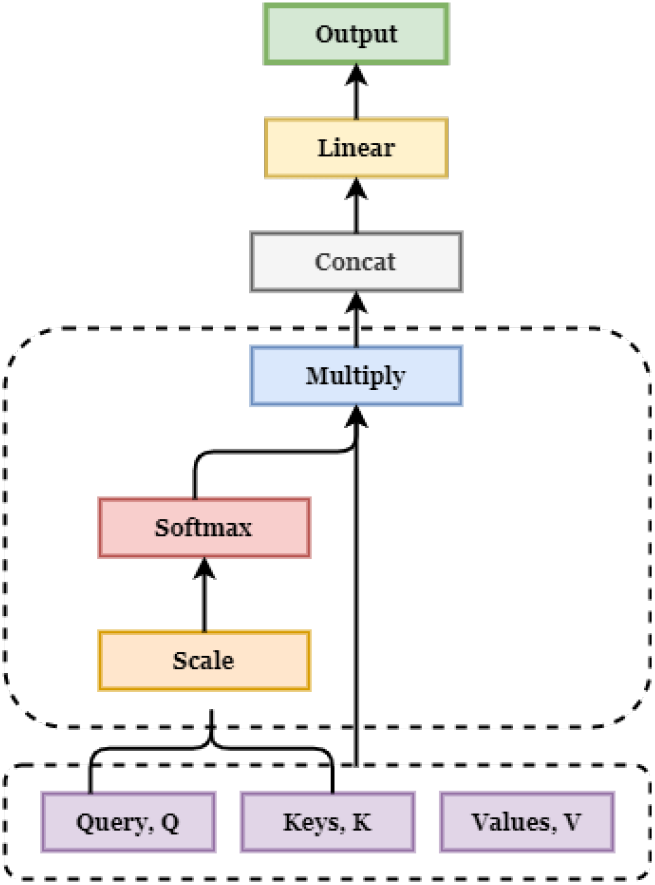
Multi-Head Attention Model.

To learn the generator’s distribution which is denoted by *P*_*g*_ over to the data *x*, an input noise denoted by *p*_*z*_(*z*) and *G*(*z*; *θg*) where a differentiable function G which represents perceptron with parameter *θg*. The *θg* is used to describe a mapping to data space. Single output scalar can be obtained by another perceptron *D*(*x*; *θd*), which has multi-layers. An initial probability obtained by the x without *P*_*g*_ is represented by *D*(*x*). Training D to assign the right label to the extensive training samples and the samples of G with a higher probability. To train G to minimize *log*(1 − *D*(*G*(*z*))) at the same time [32 - 36]:

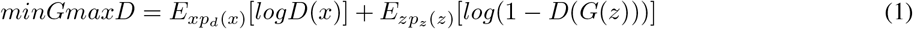

Because of some advantages over traditional deep generative models (DGMs), GANs, is also a common family member of DGMs, have gained exponentially expanding attention in the field of deep learning. GANs are more capable of producing output than other DGMs. When compared to the most well-known DGM, the variational autoencoder (VAE), GANs may generate any form of probability density, whereas VAE cannot generate clear and sharper images. The latent variables can be any size without any restrictions [33]. However, GANs have achieved state-of-the-art performance in synthesizing and generating data. In this experiment we have utilized GAN for ECG data synthesis.

### 3.2 Overview of Attention Model

Attention Model (AM), which was first proposed for Machine Translation, has now become a widely used concept in neural network research. Within the Artificial Intelligence (AI) field, attention has exploded in popularity as a crucial component of neural architectures for a staggering variety of applications in Natural Language Processing (NLP), Speech recognition, and Computer Vision (CV) [9]. Attention model is introduced based on encoder-decoder architecture.

An attention concept defines the conversion of a query and a collection of key-value pairs to an output, where each components are considered as vectors such as key, value and query. Key is responsible to determine the query’s compatibility function and the result is defined by the weighted sum of all values [26 - 28]. Scalar dot product is composed as an attention model. Dimensions of key, query and values *d*_*k*_, *d*_*v*_ can reconstruct the output. Dot products are computed of queries with corresponding keys to initiate the weights on each value, which is divided by 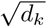 and to the end we apply a softmax function. We possibly obtain the attention function with the basis of brunch of queries which are combined into a matrix *Q*. The matrices such as *K* and *V* are denoted for key and value respectively which are also packed together. The output matrix is calculated as follows:

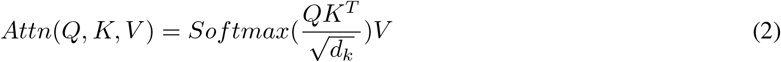

Additive and dot-product (multiplicative) attention are the two most widely employed attention functions. The scaling factor of 1*/d*_*k*_ is excepted, dot-product attention is similar to our approach. By single hidden layer a feed-forward network is constructed, additive attention calculates the compatibility function. While the theoretical complexity of the two is similar, dot-product attention is more complicated. Since it can be done using highly optimized matrix multiplication code, it is significantly quicker and more space-efficient in practice. While both techniques perform equally for small values of *d*_*k*_, additive attention beats dot product attention without scaling for bigger values of *d*_*k*_. We believe that when *d*_*k*_ increases, the size of the dot products increases, pushing the softmax function into areas with extremely minor gradients. To overcome this, we increase the size of the dot products by a 1*/d*_*k*_ [9, 26, 30].

### 3.3 Overview of Multi-Head Attention Model

Multi-head Attention is an attention mechanism module that cycles through an attention mechanism several times in parallel. After that, the separate attention outputs are combined and linearly converted into the anticipated dimension. Multiple attention heads, on the surface, appear to allow for various approaches to different portions of the sequence.

We observed that linearly determining the values with *h* times with distinct, keys and queries, and they learn from the projection with dimensions *d*_*q*_, *d*_*k*_, and *d*_*v*_, and then a single attention function with having *d*_*model*_ with a dimensional keys, queries and values. Parallel operation of attention function on individual of all are projected versions of Q, K and V (Queries, keys and values) with expecting *d*_*v*_-dimensional output values. These would be concatenated and projected again, yielding the final values, as illustrated in Figure 4. Finally, the results are concatenated to form the final output. In the center, linear transformations are applied successively. The entire formulation is written as follows [26, 30]:

**Figure 4:**
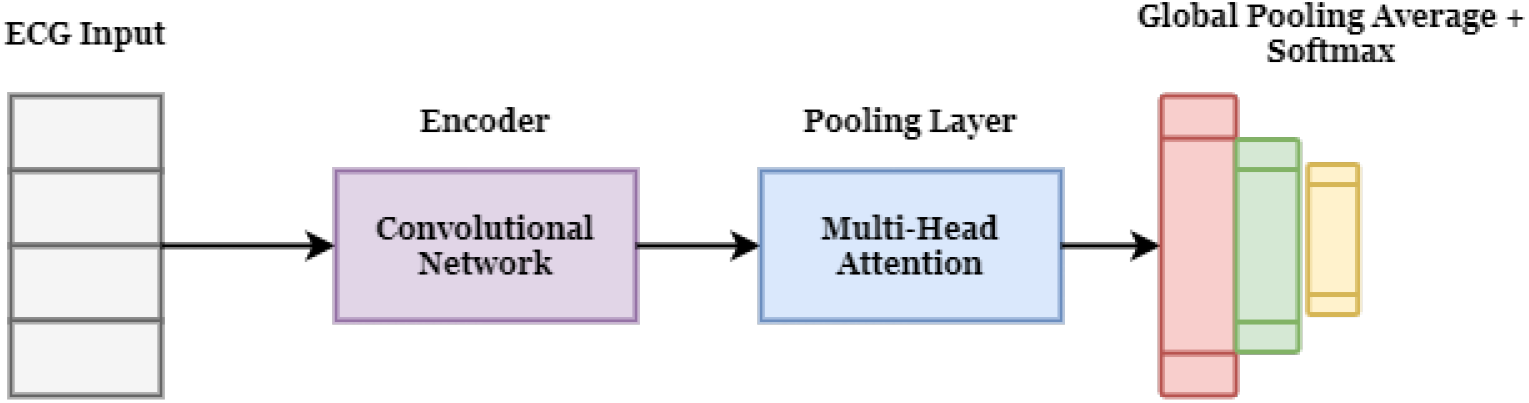
*Proposed HeartNet* Model.

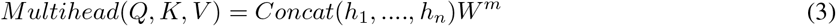

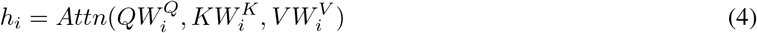

Where, 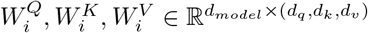. Multi-head attention allows the model to simultaneously attend to input from several representation subspaces at various locations. Averaging prevents this with a single attention head.

### 3.4 Convolutional Network

Convolutional Network or CNN algorithm is based on deep learning. CNN is quite well-known in the area of deep learning based computer vision and image processing as an extensively used technique. Generally, it is made up with three layers such as an *i* layer (input), *o* layer (output), and the *h* layer (hidden). Hidden layers are the most essential part of an CNN model. It is also known as intermediate layers. These layers are disguised by activation function, pooling layer and convolution. Convolution layer mainly reduces the size of an input and diminish the overall computation. However, the overall structure of an CNN model is constructed with input layer, convolution layer, pooling layer, fully-connected layer, and softmax layer. Convolution and pooling layers, unlike standard neural networks, can extract and map features from input data to speed up learning and decrease over-fitting. Because the CNN has a layered perception characteristic, 1D and 2D CNNs are widely utilized [17].

### 3.5 Proposed HeartNet Model

Our proposed **HeartNet** model is a combination of convolutional network model with multi-head attention mechanism. To get the proposed model intuition, we need to show a stable mathematical relation between convolution and multi-head attention. Convolutional layers are the unilateral method for building image-processing neural networks.

#### Multi-Head Self Attention and Convolutional Layer

To see how a convolutional neural layer and a self-attention layer are similar and different, consider how each of them processes an image of the shape *W* × *H* × *C*_*in*_. Although the processes of CNNs are widely understood, moving the transformer architecture from 1D to 2D needs a thorough understanding of self-attention dynamics. As we know a CNN model has several convolutional layers and subsampling layers. Let the kernal size of the filters are *k* × *k*. And the dimensions the input and output are expressed as *C*_*in*_ and *C*_*out*_. And a bias vector of *b* can be considered. A key/query size *D*_*k*_, a head size *D*_*h*_, a number of heads *N*_*h*_, and an output dimension *D*_out_ constitute a self attention layer. A key matrix 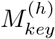, a query matrix 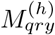, and a value matrix 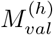 for each head *h*, as well as a projection matrix *M*_*out*_ used to put all heads together, parameterize the layer [31].

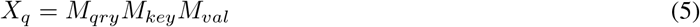

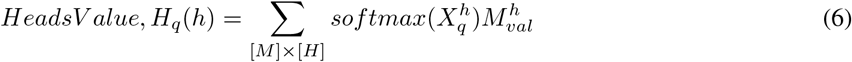

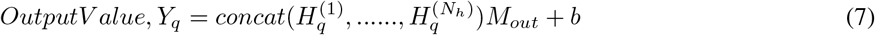

Figure 4 illustrates our proposed *HeartNet* model. We utilized ECG input then feed into the convolutional block and that block worked as an encoder. Then instead of the pooling layer we have utilized the multi-head attention unit. We have considered the number of heads to 4 and 8 in our experiment. However, we also utilized global pooling average instead of fully-connected layer and softmax in an output layer. A summary of our proposed model is given in Table 2.

**Table 2:**
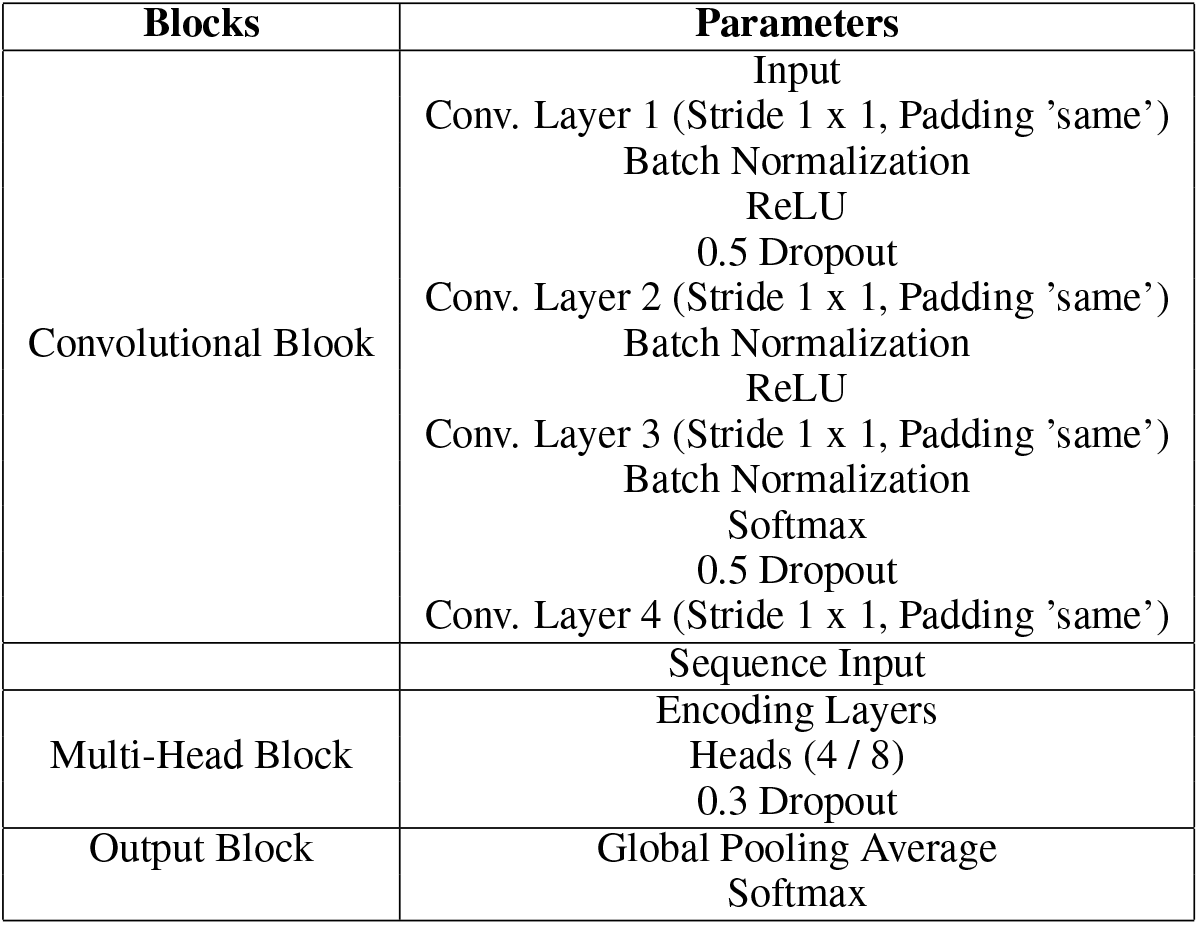
Summary of Our Proposed Model.

## 4 Experiments

### 4.1 Datasets

#### 4.1.1 MIT-BIH Dataset

In this experiment, we have derived an open-sourced dataset named MIT-BIH [11]. The original ECG signals are noted by more than two cardiologists in every record. This dataset comprises 48 ECG recordings, each of which has a 30-minute segment chosen from 48 individuals’ 24-hour records. Each ECG signal is captured at 360 Hz after passing through a band pass filter at 0.1–100 Hz. There are only 44 records are used in this experiment from the MIT-BIH dataset. Arrhythmias of many sorts may be found in this dataset. However, there are 15 different sub-categories under 5 categories. These 15 types of arrhythmia are fall under N, S, V, F, Q. Where N refers for normal heartbeat, Q refers for unknown heartbeat, V refers for ventricular ectopic heartbeat, F refers for fusion heartbeat and S refers for ventricular ectopic heartbeat. A holistic overview of the dataset with 15 different sub-categorical description are shown in Figure 5.

**Figure 5:**
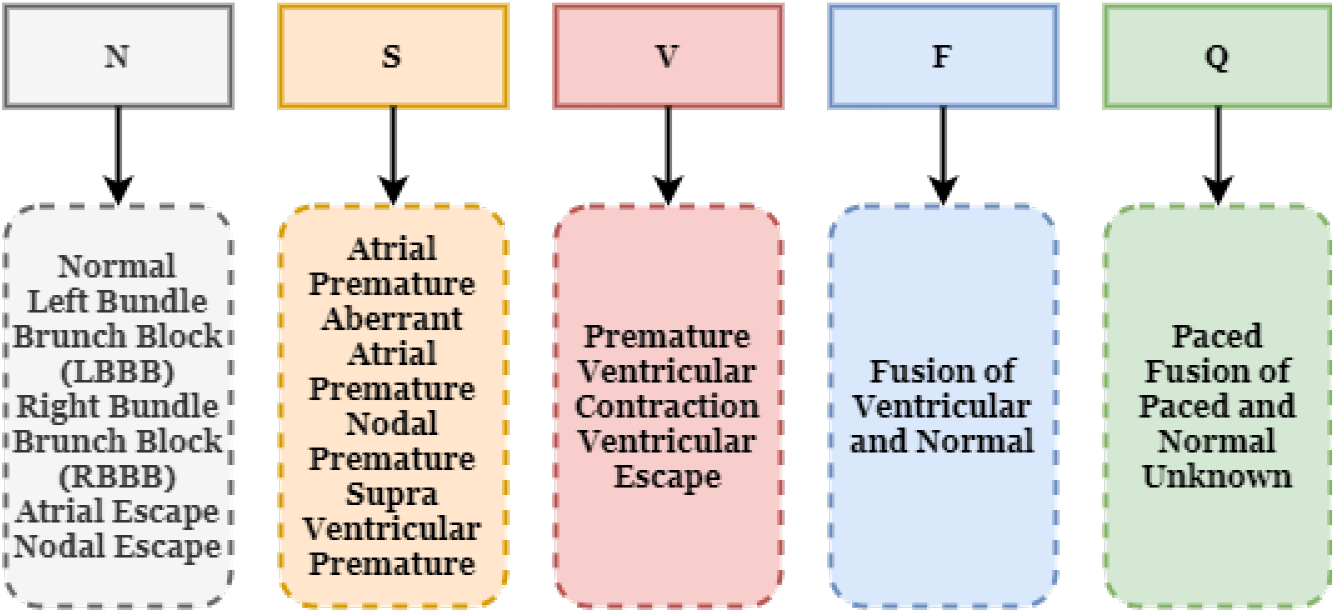
15 Sub-Categories Description from the Dataset.

#### 4.1.2 Atrial Fibrillation Dataset

A recent dataset called Atrial Fibrillation Dataset [39] is used in our experiment. Which was published in 2017 in a PhysioNet/CinC challenge. This dataset contains four classes such as normal, artrial fibrillation (AF), noisy and unclassified. There are total 8526 recordings whereas 5154 recordings are normal, 771 recordings are atrial fibrillation class, 2557 recordings are unclassified and 46 recordings are noisy.

#### 4.1.3 PTB Dataset

Another dataset is used in our experiment called Physionet PTB Diagnostic Database [39]. It has around 550 recordings of 290 subjects. Where 209 subjects are men and 81 subjects are women. Where 148 diagnosed as myocardial infraction (MI), 52 normal control and 7 associated with different diseases such as myocardial infarction (MI), Heart failure, branch block, dysrhythmia, myocardial hypertrophy, valvular heart disease etc.

### 4.2 Data Synthesis with GAN

Medical data synthesis is considered as fabricated data closer to the original patient record. It is not the same as partially counter data or data sets with variables filtered or deleted in order to limit protected health information factors. Data synthesis is needed to validate machine learning performance in larger balanced datasets. In this experiment, we have also adopted data synthesis utilizing state-of-art GAN technique.

In the previous sub-section we have discussed about the dataset. However, there are five individual categories in the dataset such as N, S, V, F, Q. In the N (Normal) category there are 90589 data, Q (Fusion of paced and normal) has 8039 data, V (Premature ventricular contraction) has 7239 data, S (Atrial premature) has 2779 data and F (Fusion of ventricular and normal) has 803 data. So we can derive that the dataset is imbalanced. As the S and F have a very less data points, so GAN is adapted to increase the number of data points in S and F. After acclimating GAN, number of data points in S and F have been enhanced. Now S has 5179 data and F has 2339 data. The main objective of adapting GAN is to validate the robustness of the developed model with slightly higher number of training data in each class. In the GAN training, we have set epochs to 3000 and observed the training in every 300 epochs interval. We have also observed the discriminator (D) loss and generator (G) loss in respect to fluctuating time frame. In the Figure 6 GAN training graphs are shown. And Figure 8 represents the generated synthesized data sample for F category.

**Figure 6:**
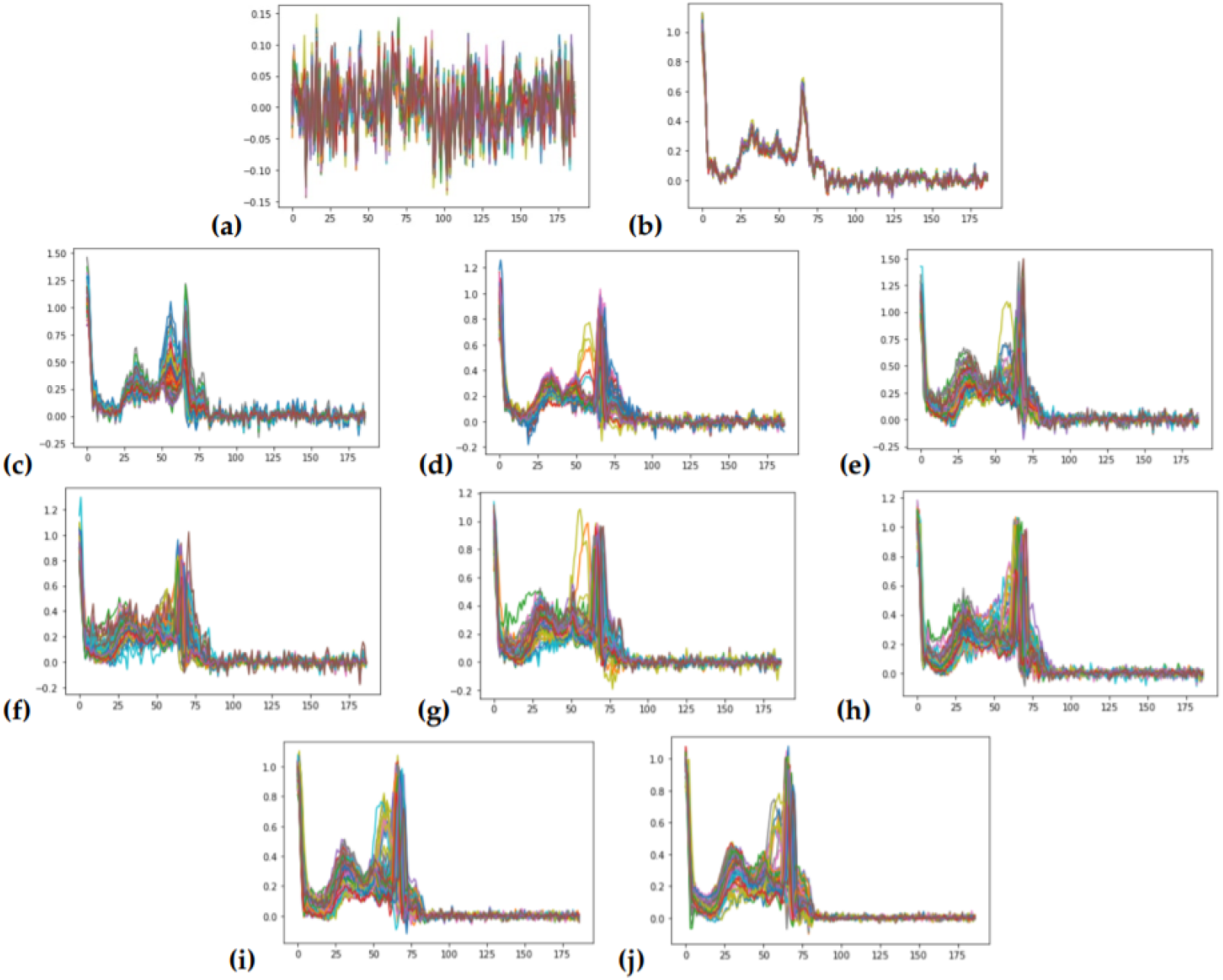
Training Graphs of GAN for Data Synthesis in 3000 Epochs. (a) Epoch: 300 | Loss_D: 1.35 | Loss_G: 0.71 | Time: 00:49:58, (b) Epoch: 600 | Loss_D: 0.37 | Loss_G: 2.4 | Time: 00:56:06, (c) Epoch: 900 | Loss_D: 0.21 | Loss_G: 2.6 | Time: 01:01:54, (d) Epoch: 1200 | Loss_D: 0.55 | Loss_G: 2.34 | Time: 01:07:34, (e) Epoch: 1500 | Loss_D: 0.94 | Loss_G: 1.05 | Time: 01:13:48, (f) Epoch: 1800 | Loss_D: 0.88 | Loss_G: 1.48 | Time: 01:20:19, (g) Epoch: 2100 | Loss_D: 0.62 | Loss_G: 1.51 | Time: 01:26:40, (h) Epoch: 2400 | Loss_D: 0.90 | Loss_G: 1.07 | Time: 01:34:01, (h) Epoch: 2700 | Loss_D: 2.16 | Loss_G: 1.51 | Time: 01:41:32, (h) Epoch: 3000 | Loss_D: 0.87 | Loss_G: 1.76 | Time: 01:47:01.

**Figure 7:**
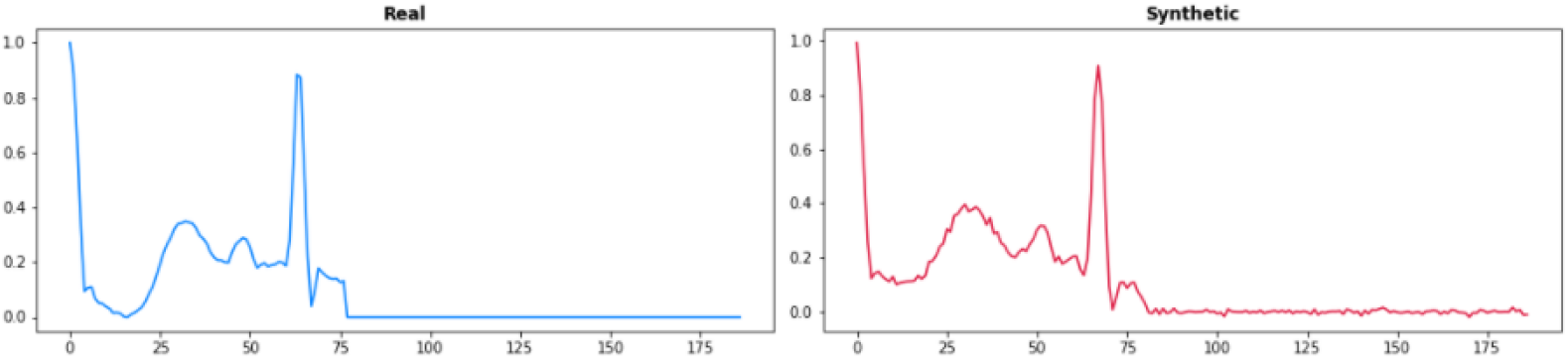
Fusion of Ventricular and Normal (F) Category Data Synthesis with GAN Output.

**Figure 8:**
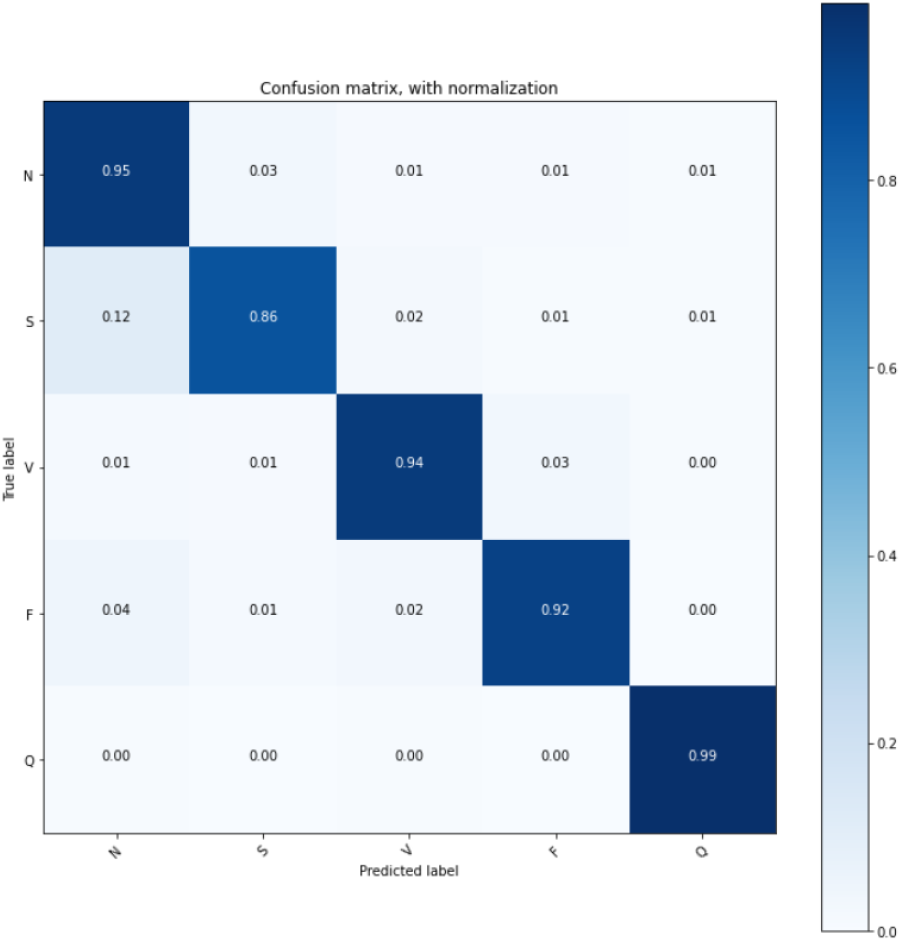
Confusion Matrix of Our Proposed Model on MIT-BIH Database.

### 4.3 Evaluation Matrix

Our experiment is a multi-class classification problem. Considering the guidelines of AAMI, there are few extensive evaluation techniques are widely utilized such as Accuracy, Precision, Recall and F1-Score. Where accuracy compromises the overall proportion of correctly predicted labels. Precision mainly denotes the number of points correctly predicted out of all positive predicted classes. Recall denotes the number of points correctly classified out of all samples. F1-score is the weighted average of precision and recall. MCC becomes popular in imbalanced cases, which denotes a correlation between predicted class and true class. Whereas specificity is an alternate version of recall which signifies the rate of correctly classified negative samples.

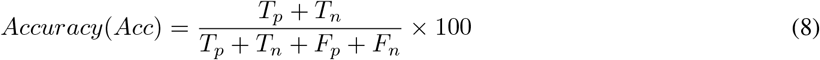

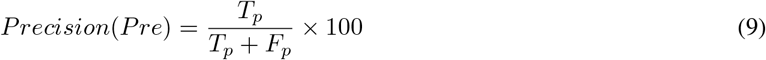

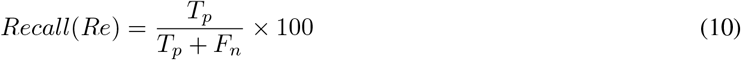

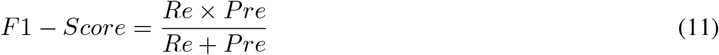

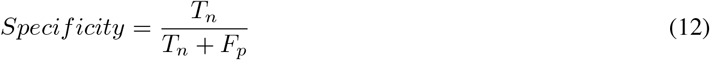

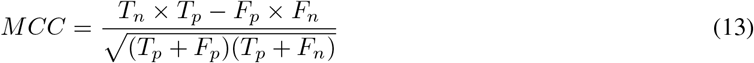

Here,

*T*_*p*_ = True Positive

*T*_*n*_ = True Negative

*F*_*p*_ = False Positive

*F*_*n*_ = False Negative

### 4.4 Experimental Results and Discussions

Initially, we worked with MIT-BIH database for arrhythmia classification. We set up five individual categories on arrhythmia classification task. At first we utilized two different datasets to see performance on those dataset of our proposed model. One dataset is synthesized with GAN and another one is not synthesized. However, we also utilized one of the very recent datasets on atrial fibrillation detection which has four classes such as normal, atrial fibrillation, unclassified and noisy. As we can see from the Table 3 that our proposed model has obtained 87.29 ± 2.42 prediction accuracy and 86.96 ± 1.98 precision in the normal category. Where normal class has the higher number of data samples considering other categories. On the other hand, in the atrial fibrillation (AF) category our proposed model obtained 85.16 ± 1.79 prediction accuracy and 83.14 ± 1.14 precision. However, in unclassified and noisy category, our proposed model has achieved 81.96 ± 2.43 and 76.21 ± 1.39 prediction accuracy respectively. Full performance report including MCC and specificity on the atrial fibrillation database is shown in Table 3. And another database has two categories such as normal and abnormal. However, the abnormal category has various sub-classes such as myocardial infarction, bundle branch block, arrhythmia, cardiomyopathy/heart failure, myocardial hypertrophy, etc. In our experiment, our developed model has performed significantly well with a prediction accuracy and precision of 98.72 ± 1.14 and 97.82 ± 1.01 respectively. Table 4 demonstrate the experimental results on PTB database on each class with detailed outcomes.

**Table 3:** Our Proposed Model’s Performance on Atrial Fibrillation Database.

| Category | Accuracy (%) | Precision (%) | F1-Score (%) | Recall (%) | Specificity (%) | MCC (%) |
| --- | --- | --- | --- | --- | --- | --- |
| Normal | 87.29 ± 2.42 | 86.96 ± 1.98 | 88.19 ± 1.76 | 89.76 ± 1.74 | 87.61 ± 2.07 | 67.46 ± 1.72 |
| AF | 85.16 ± 1.79 | 83.14 ± 1.14 | 86.42 ± 1.69 | 84.07 ± 2.42 | 85.43 ± 1.49 | 65.76 ± 1.59 |
| Unclassified | 81.96 ± 2.43 | 80.11 ± 2.11 | 83.52 ± 2.07 | 86.68 ± 1.78 | 83.36 ± 1.82 | 66.07 ± 1.80 |
| Noisy | 76.21 ± 1.39 | 77.36 ± 2.18 | 79.19 ± 1.92 | 80.13 ± 2.06 | 81.15 ± 1.46 | 61.26 ± 1.45 |

**Table 4:** Our Proposed Model’s Performance on PTB Database.

| Category | Accuracy (%) | Precision (%) | F1-Score (%) | Recall (%) | Specificity (%) | MCC (%) |
| --- | --- | --- | --- | --- | --- | --- |
| Normal | 98.72 ± 1.14 | 97.82 ± 1.01 | 98.11 ± 0.82 | 97.45 ± 1.26 | 96.70 ± 2.07 | 82.46 ± 2.11 |
| Abnormal | 97.59 ± 0.82 | 96.80 ± 1.70 | 97.48 ± 1.26 | 96.26 ± 1.92 | 94.16 ± 1.96 | 79.61 ± 2.76 |

In Table 5, we illustrated the overall performance of our proposed model without considering each category. Our proposed method has obtained 99.67 ± 0.11 prediction accuracy and 99.27 ± 0.17 precision respectively. Our proposed model also obtained 89.24 ± 1.71 MCC, which indicates the relation between predicted class and true class. Our models is constructed into four parts such as input layer, CNN blocks, Multi-head attention layer and classification layer or output layer. For the proposed model, we have utilized Adam optimizer for training with an initial learning rate of 0.001which is ideal in most cases. Dropout is set to 0.5 in CNN block. And the batch size is set to 128 for training all experiments. We tested the batch size with 32 and 64, but the 128 has be best output among three. We defined the head number is 4 and 8. Where the experiment with 8 head exhibits the best outcome in our case. We have divided the synthesized dataset into 80% and 20% for training and testing respectively. For each experiment, the epoch is set to 100. And categorical cross-entropy is used as a loss function for the proposed model. Which is generally used in multi-class classification cases.

**Table 5:** Our Proposed Model Performance on Synthesized Test Data without Considering Individual Categories.

| Model | Accuracy (%) | Precision (%) | F1-Score (%) | Recall (%) | Specificity (%) | MCC (%) |
| --- | --- | --- | --- | --- | --- | --- |
| **Proposed** | 99.67 ± 0.11 | 99.27 ± 0.17 | 98.70 ± 0.29 | 98.93 ± 0.46 | 99.16 ± 0.19 | 89.24 ± 1.71 |

At first, for the MIT-BIH database, we trained our model without synthesized data. We can see the result in Table 6, in the insufficient categories such as S and F. In supraventrical ectopic beats (S) category there are only 2779 data samples, our proposed model achieved 78.46 ± 1.97 prediction accuracy and 79.82 ± 1.44 precision. In the fusion beats F category, we have 803 data sample, which is quite insufficient to train a model in a large scale. thus, our model only manage to obtain 71.76±1.92 accuracy and 74.21±2.09 precision with a MCC of 57.52±2.46.

**Table 6:**
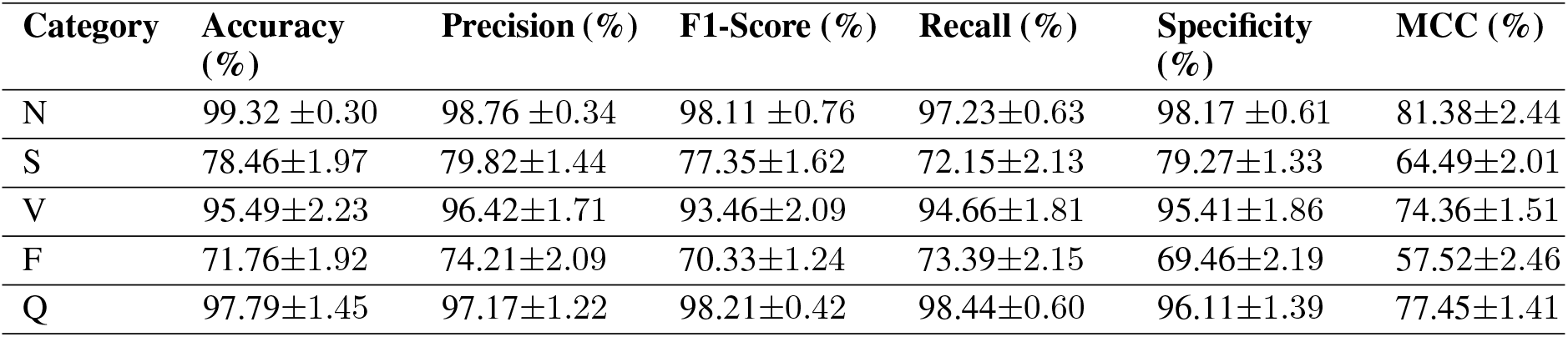
Our Proposed Model Performance without GAN-based Synthesized Data.

| Category | Accuracy (%) | Precision (%) | F1-Score (%) | Recall (%) | Specificity (%) | MCC (%) |
| --- | --- | --- | --- | --- | --- | --- |
| N | 99.32 ± 0.30 | 98.76 ± 0.34 | 98.11 ± 0.76 | 97.23 ± 0.63 | 98.17 ± 0.61 | 81.38 ± 2.44 |
| S | 78.46 ± 1.97 | 79.82 ± 1.44 | 77.35 ± 1.62 | 72.15 ± 2.13 | 79.27 ± 1.33 | 64.49 ± 2.01 |
| V | 95.49 ± 2.23 | 96.42 ± 1.71 | 93.46 ± 2.09 | 94.66 ± 1.81 | 95.41 ± 1.86 | 74.36 ± 1.51 |
| F | 71.76 ± 1.92 | 74.21 ± 2.09 | 70.33 ± 1.24 | 73.39 ± 2.15 | 69.46 ± 2.19 | 57.52 ± 2.46 |
| Q | 97.79 ± 1.45 | 97.17 ± 1.22 | 98.21 ± 0.42 | 98.44 ± 0.60 | 96.11 ± 1.39 | 77.45 ± 1.41 |

In Table 7, we can see the performance of adapted GAN to synthesized insufficient data in the S and F category. It drastically increases the samples in the dataset. Previously, the supraventrical ectopic beats (S) category has 2779 samples. But after applying GAN based synthesis the number of samples in S category turned out 5179. And F category had only 803 samples. Then after synthesis it has 2339 data samples. Our proposed model obtained higher accuracy and precision of 83.71 ± 1.72 and 86.23 ± 1.94 on the S category. Around 5% performance increase from our previous observation with data synthesis. On the other hand, in the F category, proposed model obtained 81.36 ± 1.89 of accuracy and 86.41 ± 2.24 of precision. There are around 10% improvement from previous observation. However, F has overall lower performance due to its insufficient data sample even after GAN-based data synthesis. Fortunately, precision of F category is marginally acceptable.

**Table 7:** Our Proposed Model Performance on Each Category with GAN-based Synthesized Data.

| Category | Accuracy (%) | Precision (%) | F1-Score (%) | Recall (%) | Specificity (%) | MCC (%) |
| --- | --- | --- | --- | --- | --- | --- |
| N | 99.43 ± 0.28 | 98.26 ± 0.47 | 98.71 ± 0.62 | 98.18 ± 0.89 | 99.26 ± 0.17 | 82.46 ± 2.41 |
| S | 83.71 ± 1.72 | 86.23 ± 1.94 | 78.33 ± 1.89 | 79.26 ± 2.18 | 82.18 ± 2.36 | 69.47 ± 1.78 |
| V | 96.93 ± 2.15 | 95.46 ± 1.90 | 96.16 ± 2.10 | 95.11 ± 1.98 | 97.01 ± 1.39 | 80.55 ± 1.69 |
| F | 81.36 ± 1.89 | 86.41 ± 2.24 | 67.82 ± 2.51 | 76.55 ± 2.28 | 80.12 ± 1.64 | 57.52 ± 2.46 |
| Q | 97.74 ± 1.27 | 96.29 ± 1.34 | 98.46 ± 0.42 | 98.46 ± 0.80 | 97.47 ± 1.12 | 76.33 ± 1.39 |

Then we up-sampled the dataset and converted the dataset into a balanced one. On each category it has 20% of sample contribution. In Table 8, we can see the results on the up-sampled dataset or a balanced dataset. But the accuracy and precision on each category is significantly less than the previous experiments. But the accuracy on S category and F category are quite similar. We obtained only 81.79 ± 1.52 accuracy on the S category and 81.54 ± 1.67 accuracy on the F category in this experiment. Which is lower than our previous observations by a significant margin of at least 8-10% on average. Figure 8 illustrates confusion matrix of our proposed model.

**Table 8:**
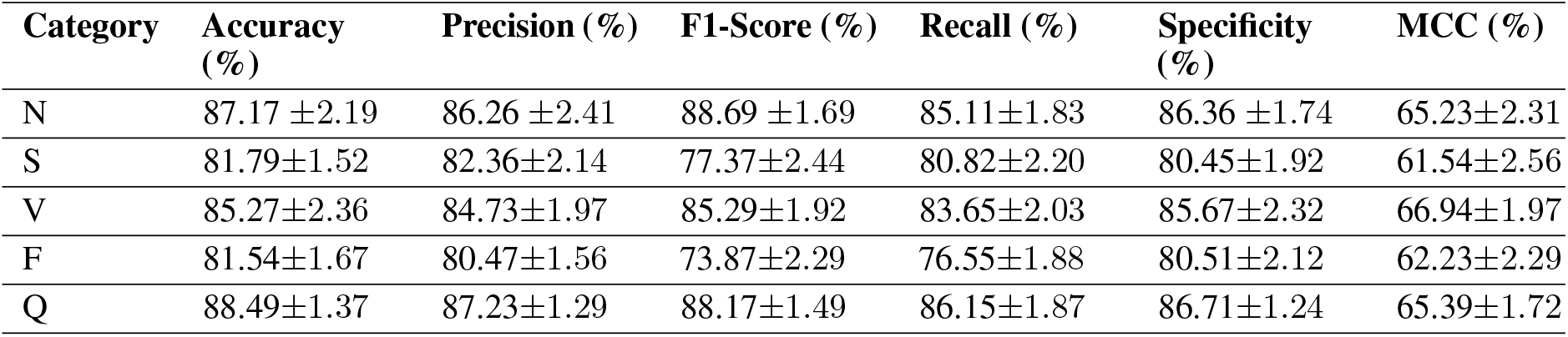
Our Proposed Model Performance with Upsampled Data.

| Category | Accuracy (%) | Precision (%) | F1-Score (%) | Recall (%) | Specificity (%) | MCC (%) |
| --- | --- | --- | --- | --- | --- | --- |
| N | 87.17 ± 2.19 | 86.26 ± 2.41 | 88.69 ± 1.69 | 85.11 ± 1.83 | 86.36 ± 1.74 | 65.23 ± 2.31 |
| S | 81.79 ± 1.52 | 82.36 ± 2.14 | 77.37 ± 2.44 | 80.82 ± 2.20 | 80.45 ± 1.92 | 61.54 ± 2.56 |
| V | 85.27 ± 2.36 | 84.73 ± 1.97 | 85.29 ± 1.92 | 83.65 ± 2.03 | 85.67 ± 2.32 | 66.94 ± 1.97 |
| F | 81.54 ± 1.67 | 80.47 ± 1.56 | 73.87 ± 2.29 | 76.55 ± 1.88 | 80.51 ± 2.12 | 62.23 ± 2.29 |
| Q | 88.49 ± 1.37 | 87.23 ± 1.29 | 88.17 ± 1.49 | 86.15 ± 1.87 | 86.71 ± 1.24 | 65.39 ± 1.72 |

### 4.5 Comparison with Other Models

In Table 9 presents the classification results in terms of accuracy by other recent works related to arrhythmia scheme. We have considered only recent works which have utilized same database and same strategy to their experiment. However, we have considered the 5-fold cross validation in our experiment, so we have considered works who did 5-fold cross validation in their experiment to compare. It allows this comparison to be fair in equal condition. Comparing to our proposed model with Abdalla [8] has marginally higher result than our approach. They utilized a CNN architecture with 11 layers construction. They worked with the same MIT-BIH dataset with a five-fold cross validation. But they did not consider the insufficient training data label problem in their experiment. Which we addressed. It has been obtained that our proposed *HeartNet* has performed higher than Kim and Pyun [10]. They utilized two different methods like bidirectional LSTM based DRNN network in their work. They also adopt preprocessing procedures like noise reduction, average filtering, etc. They run their experiment with five-fold cross validation. Kachuee et al. [16] has utilized deep CNN network with an additional residual block in their experiment. They also utilized transferable representation learning to adopt training to newer databases. Their overall accuracy is quite less comparing to other recent works.

**Table 9:**
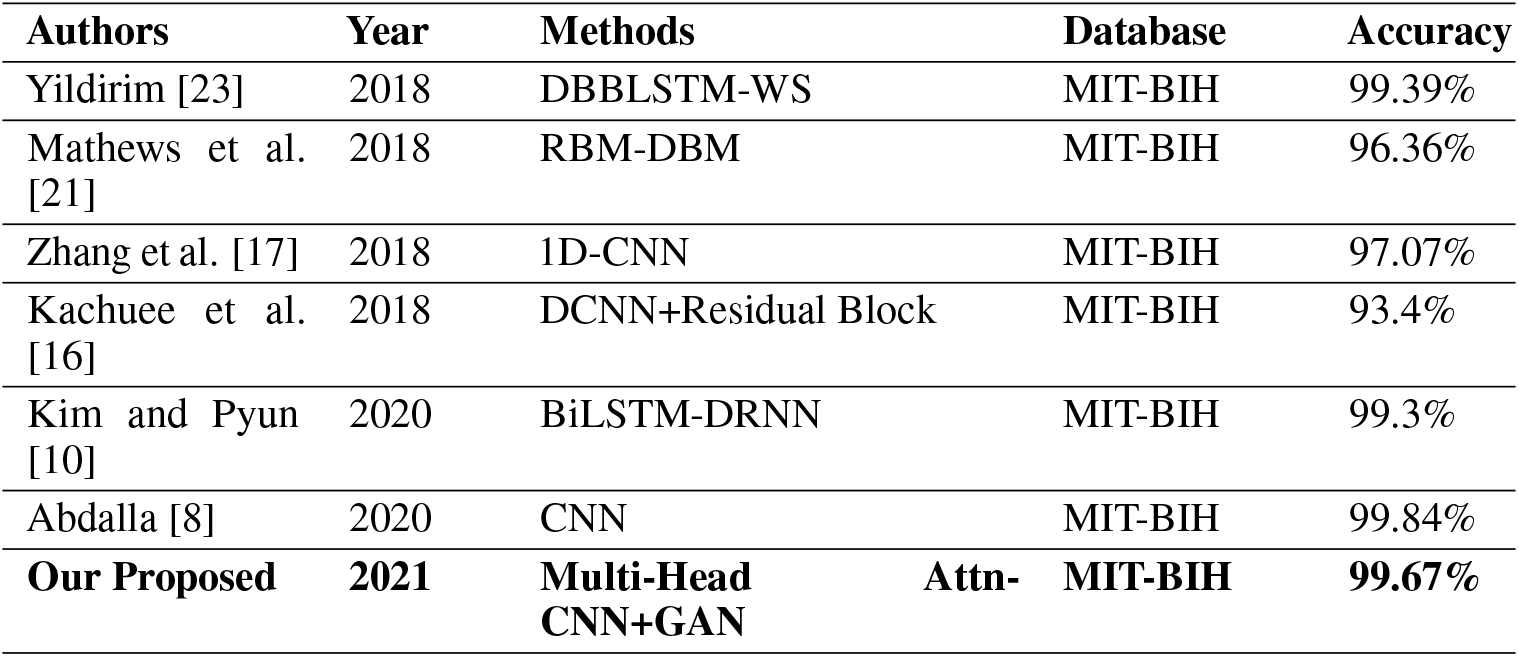
Our Proposed Model’s result compared with Other Previous Works on Arrhythmia Classification.

On the other hand, Zhang et al. [17] has achieved bare accuracy. They worked on raw ECG data with normalization before feed to their model which made of 1-dimensional CNN with fine-tuning. Other works worth mentioning like Mathews et al. [21], Yildirim [23], have also obtained very higher remarks in ECG classification on the same dataset.

## 5 Limitations of This Work

As we know, ECG recordings play a vital role to determine initial heart associated problems. Which leads to find more advanced heart problems with further diagnosis. So, an automatic ECG classifier can help the medical professionals to determine findings easily from the recorded ECGs. In this experiment, we tried to develop an automated ECG classifier with a combined deep learning technique, called HeartNet. The tried to build an effective and high-performing deep learning based ECG classifier with addressing to solve insufficient data label problem. Which can be found in recording ECG signals and build a training dataset.

Lastly, we want to discuss some difficulties and limitations that we have came across during training our model. We have trained our model in a small function in an imbalanced dataset. Where we did not observe the plateauing loss, which can lead the model to train longer. Furthermore, we have observed our model is not the best performing model on the dataset and same training settings. Our model may fail to converge its prediction if there’s too much negative samples. We further recommend to study some more advanced techniques like unsupervised or self-supervised to be applied in this domain. Because adaptation of these techniques are quite extensive that does not depend on human annotations.

## 6 Conclusion

Cardiac disease, often known as arrhythmia, is by far the most serious condition that causes mortality in humans. Doctors and medical professionals frequently use multiple-lead ECG data to detect and distinguish heartbeat signals in clinical applications. Deep learning shows to be a useful approach for dealing with the lack of labeled data that affects ECG records classification models. In recent years, various researchers have developed automatic ECG classification scheme but these are not extensively adaptable by medical professional for its less vulnerability. In this study, we proposed a new deep learning method combining multi-head attention based convolutional neural network called *HeartNet*. By utilizing convolution filters, it is quite easier to extract the features. Our primary challenge is to develop a method which can forestall the insufficient training data-label deficiency and unbalanced class distribution. To provide and overcome deficiency we adopted generative adversarial network to synthesized data and create more samples on lower distributed classes. After increasing the number of training labels, performance by our proposed model improved its accuracy by 5-10%. Our proposed model also performed well comparing to other recent models with an accuracy of 99.67% ± 0.11 and a precision of 99.27% ± 0.17. We did extensive experiment considering two other datasets to check the robustness and effectiveness of our proposed method in different scenarios. To summarize, our proposed can a confluence generic solution that could be utilized in real life applications. But we also suggest more research with some new ML techniques need to be studied.

## Data Availability

All data produced in the present work are contained in the manuscript.

## Data Availability

All data produced in the present work are contained in the manuscript.

## Acknowledgments

This research is supported by Hallym University Research Fund, 2020 (HRF-202003-016)

